# Is sobriety from alcohol necessary for evaluation of patients presenting with suicidal ideation?

**DOI:** 10.1101/2021.12.06.21267365

**Authors:** Daniel Keyes, Blake Hardin, Brandon Moore

## Abstract

**Background:** It is commonly assumed that patients intoxicated with alcohol are unreliable with respect to their statements of suicidal intent, however no prior literature evaluates the impact of sobriety on suicidal ideation (SI). In typical emergency department (ED) settings, a common practice is to wait until intoxicated suicidal individuals have reached a legally sober limit (ethanol level less than 80 mg/dL) to evaluate safety. We are not aware of any study that establishes the diagnostic reliability of the clinical suicidal ideation evaluation as a function of alcohol intoxication.

**Methods:** This study is a retrospective review of medical records for patients evaluated in a pre-COVID Midwestern ED for one calendar year. Cases were generated for review based on criteria of having a Psychiatric SW (Social Work) consult and blood alcohol level drawn while in the ED on every Wednesday and Friday of 2017 which produced 1084 cases for review. Chi-square analysis was used for comparison of variables of suicidal ideation with or without alcohol intoxication as defined by blood alcohol level (BAL) ≥80 mg/dL.

**Results:** In reference to our initial hypothesis, patients presenting with suicidal ideation and concurrent alcohol intoxication were no longer reporting suicidal ideation at sobriety in 69% of cases, compared to 38% for patients without alcohol levels on presentation. Chi-square analysis demonstrated p=0.000012.

**Conclusion:** The goal of the present study was to demonstrate, with empirical data, a relationship between alcohol intoxication and suicidal ideation. Our data suggests that patients presenting to the ED with complaints related to suicidal behavior who are found to have concurrent alcohol intoxication are more likely to deny suicidal ideation when sober than patients with similar presenting complaints and no alcohol intoxication.

## Introduction

Suicide is an important societal problem, and suicidal patients frequently present to the nation’s emergency departments (EDs). Data from the National Hospital Ambulatory Medical Care Survey reveals that there are more than 400,000 ED visits annually for attempted suicide (Canner et al. 2018) (Doshi 2005). Of these, approximately one-third of individuals are admitted to the hospital. Suicide is the 10th most common cause of death in the US and the second most common cause in the 15-34 years age group. According to the Centers for Disease Control and Prevention (CDC), in the US there were over 47,000 deaths due to suicide in 2017 and 383,000 emergency department visits for “self-inflicted injury,” which includes self-harm with or without suicidal ideation (CDC 2017).

Acute alcohol ingestion often accompanies suicidal ideation (Cherpitel et al 2004). According to data from the National Violent Death Reporting System, alcohol was detected in nearly 36% of males and 28% of female suicide decedents (Kaplan et. al 2012). Acute alcohol use is a potent risk factor for suicidal ideation and suicide attempt (Conner, et. al 2014).

It is commonly assumed that patients cannot be definitively diagnosed as suicidal until they are clinically sober. The current norm in the emergency setting is to wait until intoxicated suicidal individuals “sober up” and then reassess them for safety (Conner et. al 2014, Simpson 2019). A nationwide survey of emergency psychiatrists and behavioral health specialists found that most use “clinical sobriety” when assessing an inebriated patient, while a large subset used a patient’s repeat blood alcohol level (BAL) as compared to a pre-specified number (Simpson S 2019). A specific alcohol level of 0.08 g/dL was adopted nationally in 1998 as an enforceable marker of intoxication for motor vehicle enforcement (NIAAA 2001), and this number is also used by many practitioners as an estimate of sobriety and decisional capacity. However, we are not aware of any study that evaluates the *reliability* of the clinical suicidal ideation evaluation as a function of sobriety. Specifically, it is important to determine if intoxicated patients, who are identified to be at risk for suicide by the emergency practitioner and/or behavioral specialists, ultimately will be more or less likely to be suicidal upon sobriety as compared to non-intoxicated subjects.

In many institutions, psychiatric consultation may be used to determine discharge of the patient. In others, the emergency practitioner determines this independently. This is particularly true in the case of suicidal ideation. Only larger tertiary-care institutions typically have specialized psychiatric emergency departments, numbering approximately 100 across the US, making real-time ED evaluations by a psychiatrist uncommon (California Health Line 2019).

The importance of identifying suicidal ideation in patients before leaving the hospital cannot be overstated. If a patient presenting with suicidal ideation is sent home prematurely, they are at risk of injuring themselves or others. This could also result in legal and financial ramifications for the practitioner and the hospital or healthcare center that releases them (The Law Offices of Skip Simpson: Attorneys and Counselors).

During the COVID-19 pandemic, increases in depression and suicide have been reported, especially in the adolescent age range, less so among adults (Santomauro, D. et al. 2021) (Yard E. et al. 2021). In addition, presentations for alcohol intoxication increased substantially during the pandemic (Keyes, Hardin et al 2021). For this reason, it may be important to examine the impact of sobriety on the diagnosis of suicidality prior to the pandemic when it can be evaluated in its more natural context.

### Goals of this Investigation

Our hypothesis is that patients with blood alcohol levels >80 mg/dL and suicidal ideation are more likely to report no suicidal ideation once they are sober, as compared to a control group of patients who present without alcohol intoxication. By extension, the study addresses the question: is it necessary to wait for sobriety before defining a patient as suicidal?

## Materials and Methods

### Study Design and Setting

This was a retrospective study of pre-COVID medical records for patients who were evaluated for suicidal ideation in the emergency department from January through December 2017 when a policy of repeat ethanol level determination was in place at the study site. This research was approved by the St Joseph Health System Institutional Review Board (IRB).

### Selection of Participants

This study was performed at a moderate-sized community hospital and trauma center, with an approximate annual emergency department volume of 50,000 patients. Adults (≥ 18 years) were included if they stated suicidal intent and had an alcohol level drawn and received a social work/behavioral health specialist evaluation in the ED on any Wednesday or Friday in 2017. Blood alcohol level (BAL) was routinely drawn on all psychiatric adult patients at this institution on arrival. A repeat et BAL ≤ 80 mg/dL was required prior to social work evaluation. An ethanol level > 80 mg/dL was defined as alcohol intoxication for the purposes of this study.

### Outcome Measures

Our primary outcome was suicidal ideation upon sobriety as determined by a social worker or psychiatrist through direct interview of the patient using Diagnostic and Statistical Manual of Mental Disorders (DSM V) criteria after sobriety was achieved (American Psychiatric Association 2013). Our secondary outcomes included the presence of co-ingestants and sex.

### Data Collection, processing, and analysis

Charts were obtained by identifying all medical records of patients who received a social work/behavioral health evaluation. An abstraction tool and data dictionary were developed. Abstractors were trained in the use of the tools. After a test of these tools on a subset of records by all of the abstractors, the tools were modified and implemented for the remaining data collection. A one-week sample was used to determine sample size, including 118 social work consults, and these were filtered for any patients that were evaluated for suicidal ideation with or without alcohol intoxication. This subset of patients was used for the calculation of a sample size requirement of approximately 1050 charts. Chi-square analysis was used to evaluate the relation of suicidal ideation with alcohol intoxication, and descriptive statistics were used for age, gender, and initial ethanol level. Statistical analysis was performed using SAS statistical software (SAS, version 9.4, SAS Institute Inc., Cary, NC).

## Results

There were a total of 1084 charts reviewed for the study (Figure 1). Participants had a mean age of 39.5 years and females constituted 48% of the participants. Of the 740 excluded, 451 did not express SI, 102 were under 18, 148 were not evaluated by ED SW (132 no ED SW, 14 community outpatient for psychiatric emergencies (COPE) patients, 2 left against medical advice (AMA), and 39 were duplicate generated consults with the same financial identification number (FIN).

**Figure 1:**
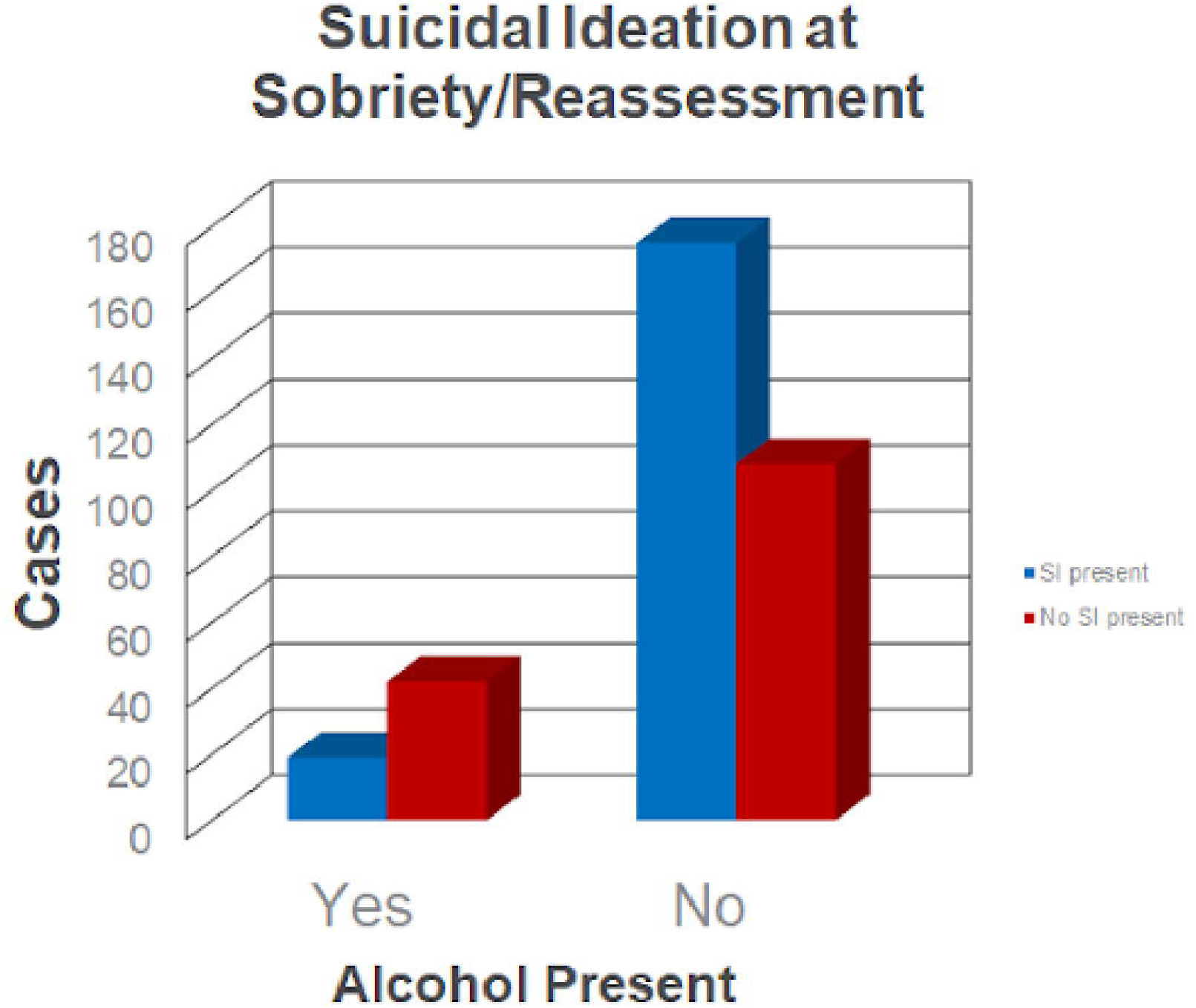
Study Flowchart. BAL = blood alcohol level; SI = suicidal ideation; SW = social worker/behavioral health specialist. FIN = financial identification number (specific to that visit). ED = emergency department. ETOH = ethanol intoxication

There were 344 total cases identified with positive suicidal ideation at initial emergency practitioner (EP) evaluation. Of these, 61 were found to have a BAL ≥ 80 mg/dL, and 19/61 (31%) continued to have suicidal ideation at sobriety, whereas 42/61 (69%) no longer expressed suicidal thoughts. In contrast, of the 283 cases with an initial BAL < 80 mg/dL, 175/283 (62%) remained with suicidal ideation at re-assessment and 108/283 (38%) no longer expressed suicidal thoughts (Chi-square p<.0001) (Table 1 and Figure 2).

**Table 1:**
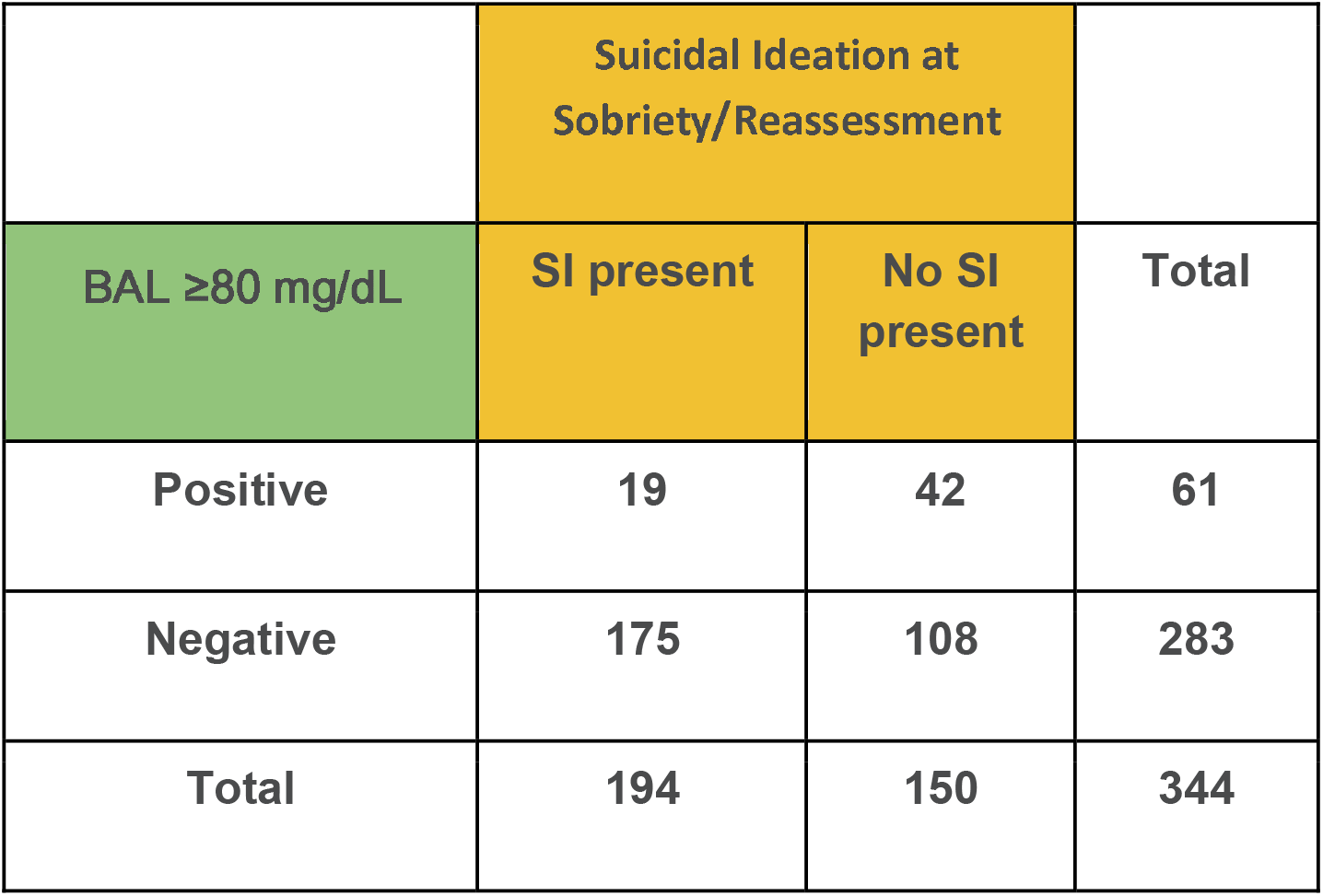
Emergency department cases in which suicidal ideation is being evaluated compared against alcohol intoxication. BAL = blood alcohol level, SI = suicidal ideation

**Figure 2:**
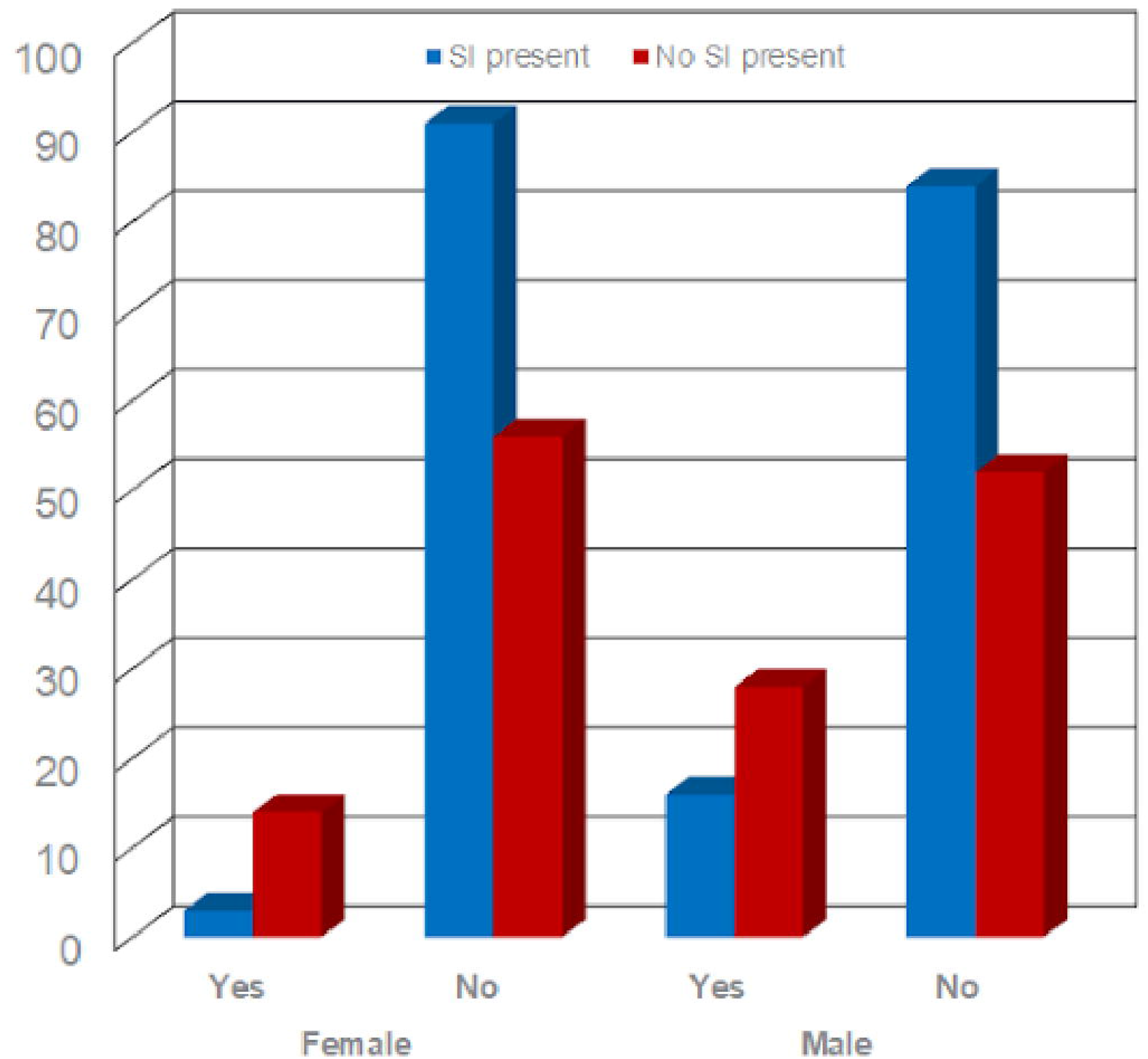
Emergency department cases in which suicidal ideation is being evaluated compared against alcohol intoxication.

This difference persisted when analyzing for individual sex, both for males (p=0.003) and females (p= 0.0005) (Table 2 and Figure 3). Alcohol intoxication upon presentation to the ED was more common in males (44/180, 24%) than in females (17/164, 10%). Stratification by race was attempted, but no significant results were obtained due to a low number of African American, Hispanic, and Asian cases. There was a significant difference within the Caucasian cohort (p<0.001). Of the 344 cases evaluated for SI, 15 cases did not have race reported.

**Table 2:**
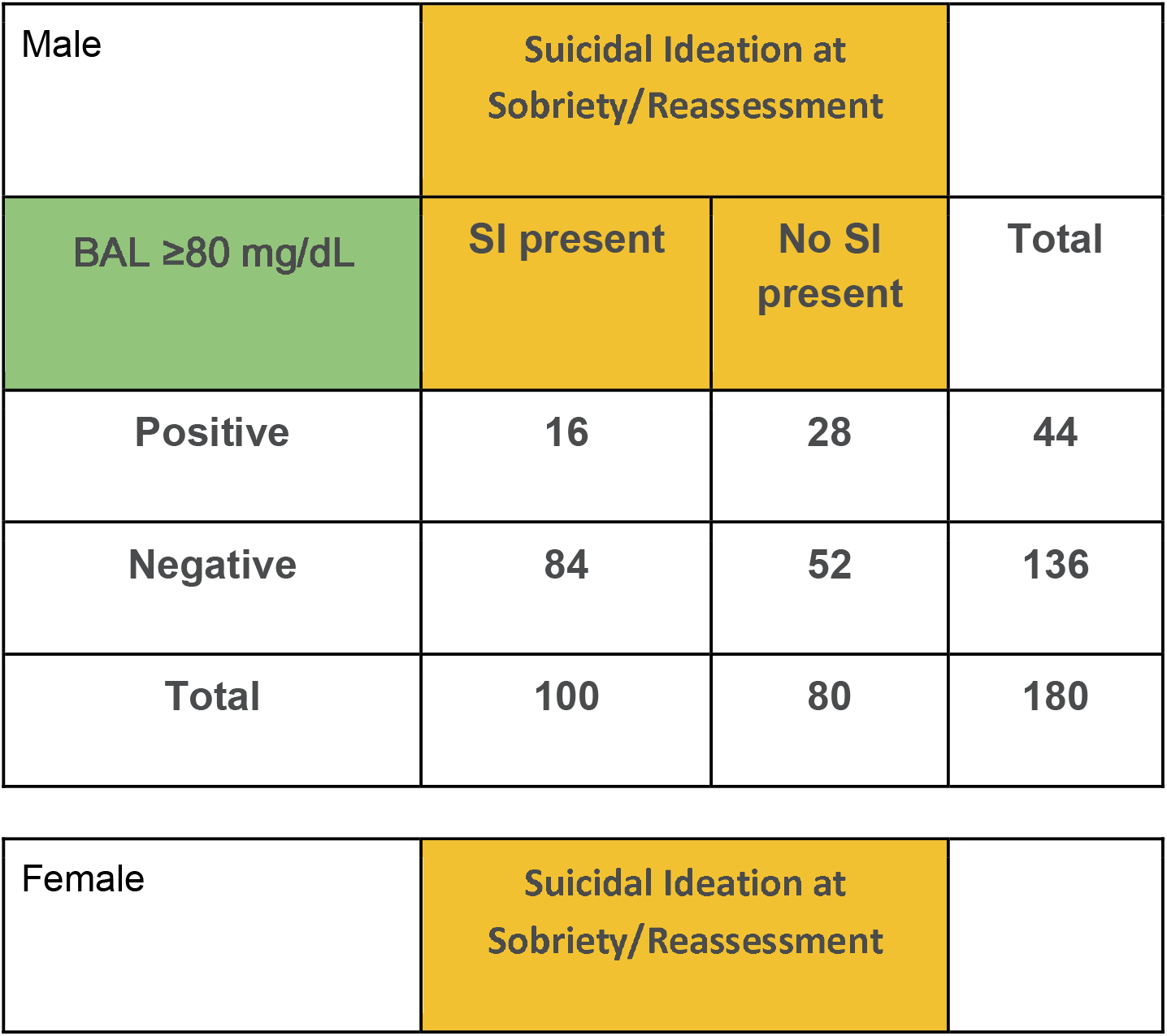
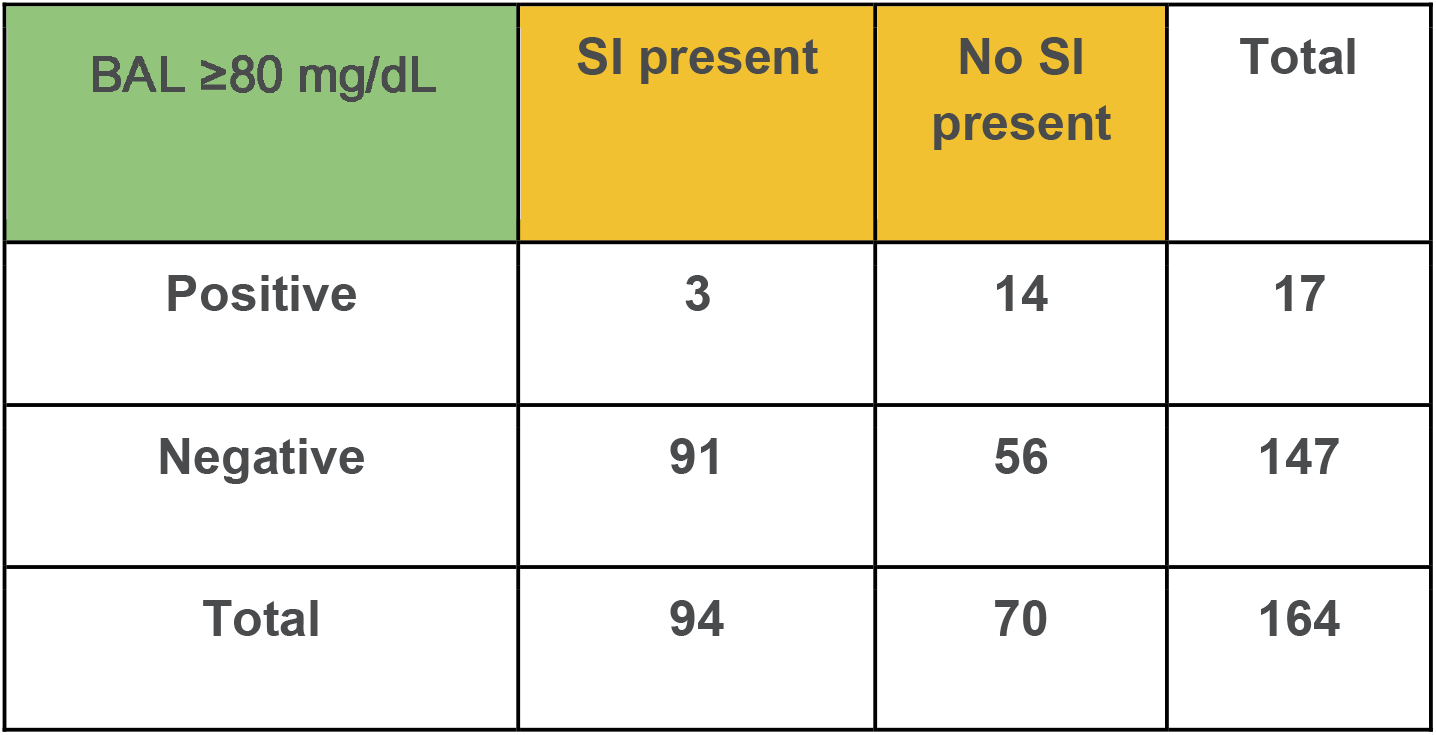
Emergency department cases in which suicidal ideation is being evaluated compared against alcohol intoxication. Gender subanalysis. BAL = blood alcohol level, SI = suicidal ideation

**Figure 3:**
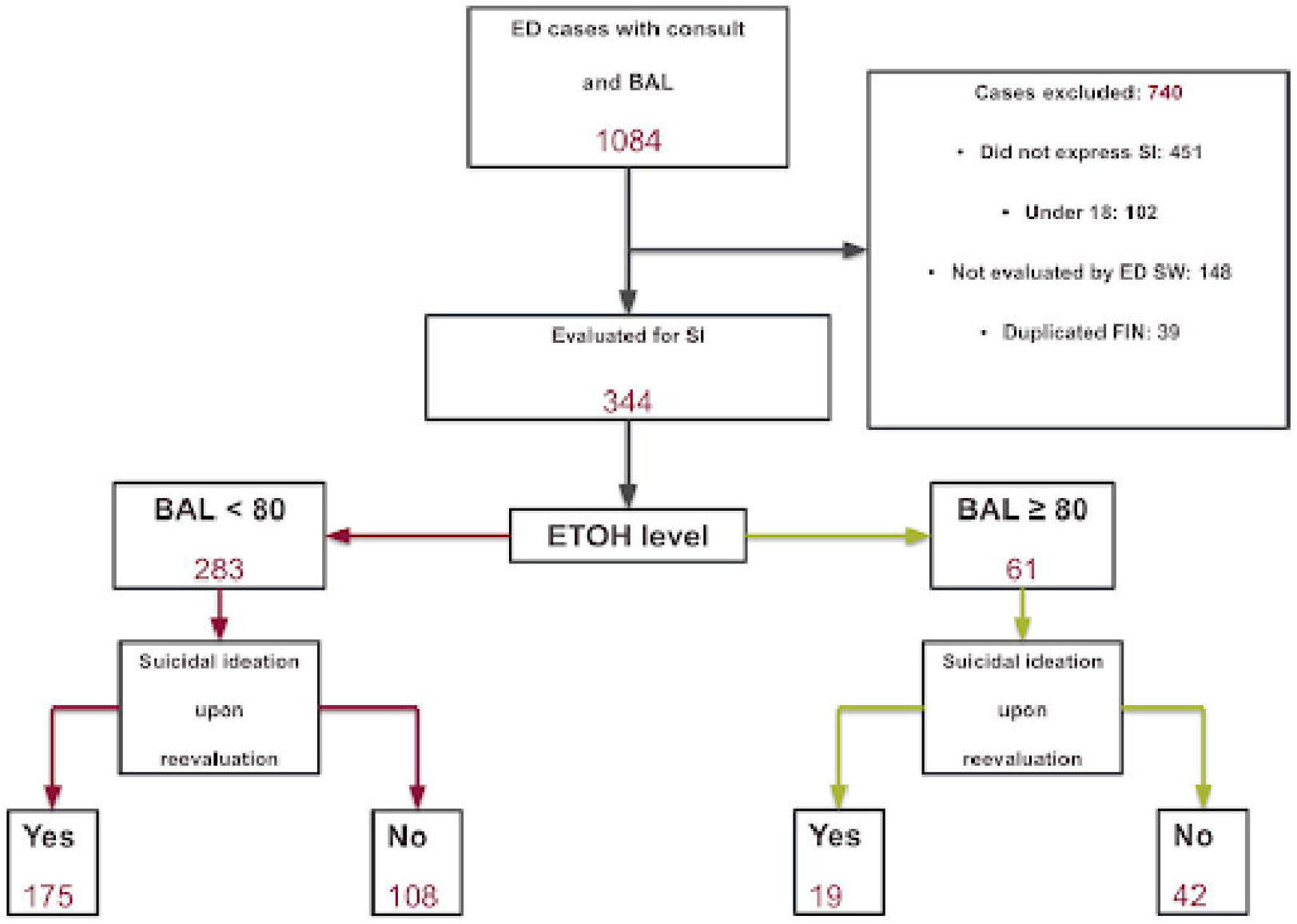
Emergency department cases in which suicidal ideation is being evaluated compared against alcohol intoxication. Gender subanalysis.

Depression was the most common psychiatric diagnosis, 230/344 (67%, Table 3). Tetrahydrocannabinol (THC) was the most common agent found in urine drug screens, found in 115/197 (58%) positive drug screens (Table 4).

**Table 3:**
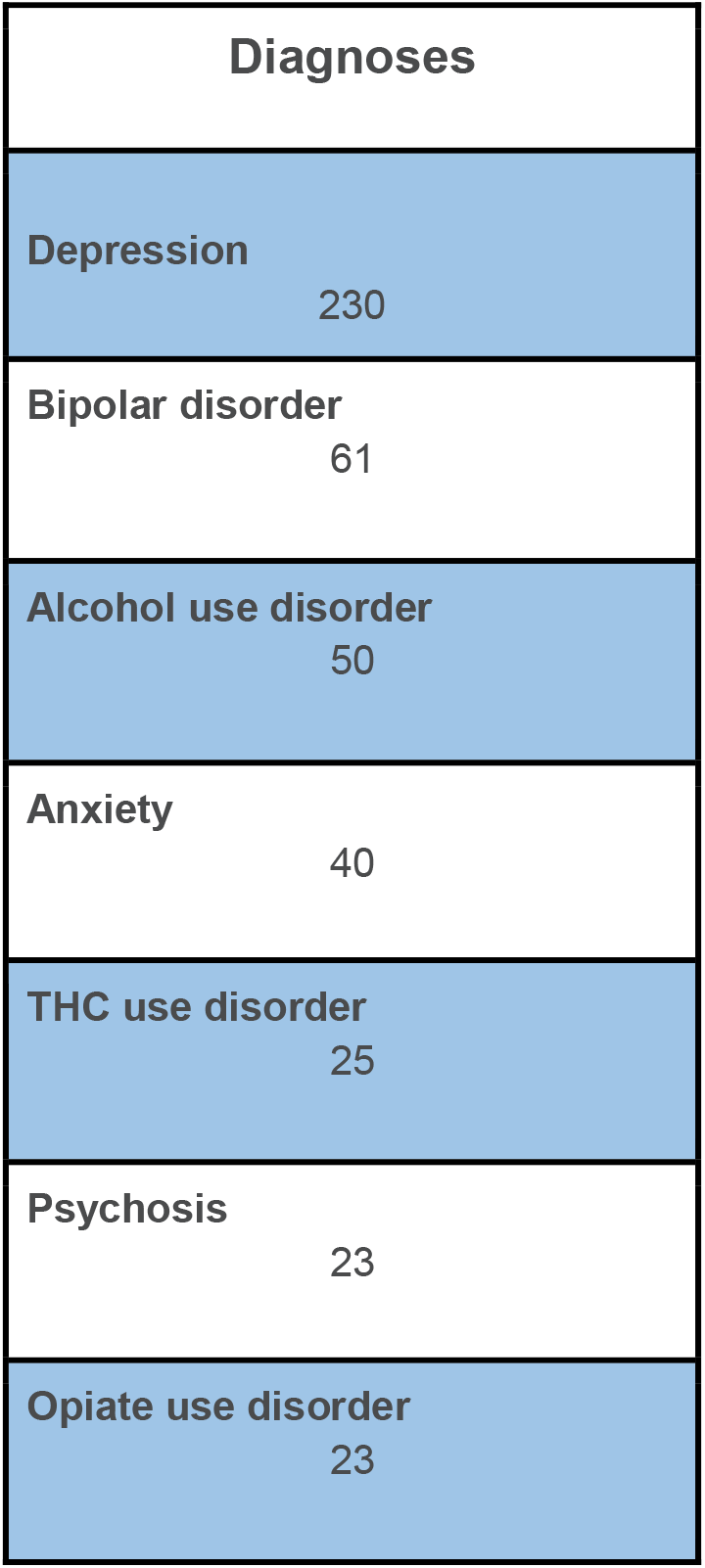
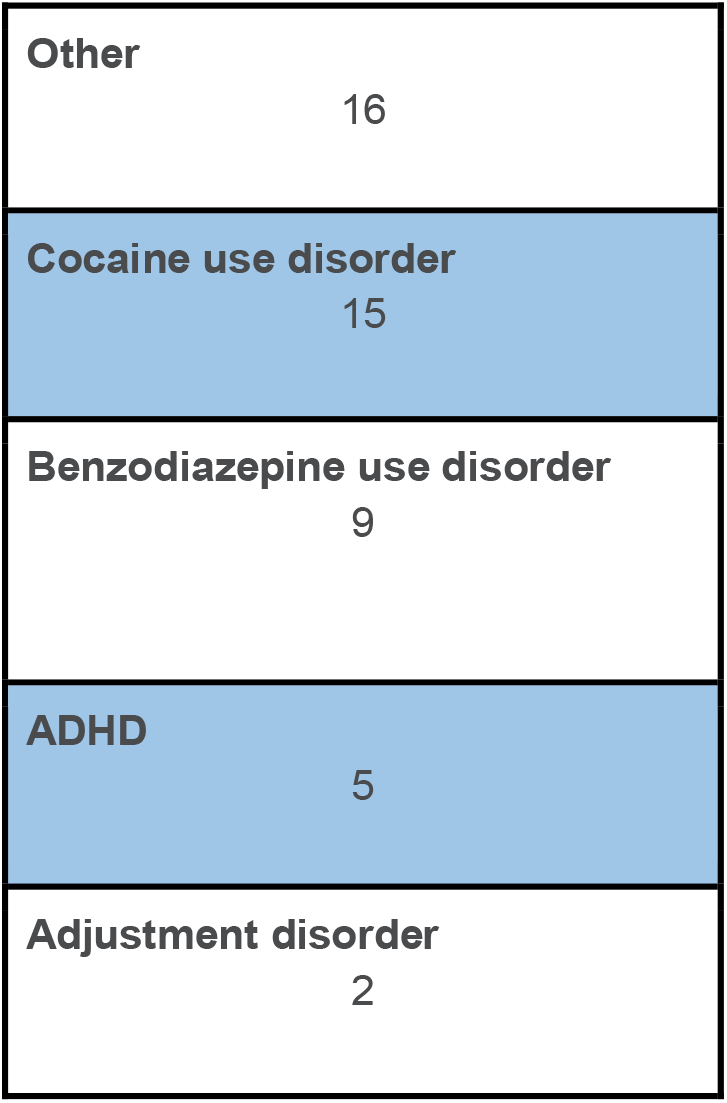
Top 12 Most Common Diagnoses Among Patients Presenting with Suicidal Ideation. The total number of patients being evaluated in the emergency department for suicidal ideation is 344. BAL = blood alcohol level, SI = suicidal ideation, THC = tetrahydrocannabinol, ADHD = attention-deficit/hyperactivity disorder

| Diagnoses |
| --- |
| Depression<br>230 |
| Bipolar disorder<br>61 |
| Alcohol use disorder<br>50 |
| Anxiety<br>40 |
| THC use disorder<br>25 |
| Psychosis<br>23 |
| Opiate use disorder<br>23 |

|  |  |
| --- | --- |
| Other | 16 |
| Cocaine use disorder | 15 |
| Benzodiazepine use disorder | 9 |
| ADHD | 5 |
| Adjustment disorder | 2 |

**Table 4:** Most common co-intoxicants done in a standard urine drug screen done in the emergency department. THC = tetrahydrocannabinol

| Co-Intoxicant | Number of Cases | Percent of Study Cases |
| --- | --- | --- |
| THC | 115 | 33.4% |
| Benzodiazepines | 92 | 26.7% |
| Cocaine | 52 | 15.1% |
| Opiates | 38 | 11.0% |
| Amphetamines | 15 | 4.4% |
| Barbiturates | 9 | 2.6% |
| No co-intoxicant | 147 | 42.7% |

## Discussion

In this retrospective study, we found that patients who were intoxicated with alcohol and expressed suicidality upon initial evaluation by the emergency practitioner were much less likely to be suicidal upon sobriety by behavioral health personnel than patients who were not initially intoxicated.

Emergency and behavioral health personnel face a challenge when evaluating patients with SI. Today, it is common to await sobriety prior to determining safety risk. Current guidelines recommend assessing the decisional capacity of the patient prior to proceeding with the full safety assessment (Allen M et al 2015). A recent survey noted that 58% of emergency psychiatrists and behavioral health specialists “proceed once the patient is clinically sober” (Simpson SA 2019). However, this widely-held assumption that evaluation of suicidality is only valid in patients who are not intoxicated has not been previously tested.

There is an inherent risk of death for patients who are discharged with suicidality. In addition, mental health inpatient resources are increasingly scarce, and it is critical that an efficient and accurate allocation of resources exist in this realm (Wood et al. 2018). The risk tolerance of individual emergency practitioners may also be influenced by medico-legal concerns when discharging a suicidal individual. These important factors result in heightened concern to accurately triage select patients to inpatient services.

Intoxication is inherently associated with high-risk behaviors, and this includes suicidal ideation and attempted suicide (Darvishi et al. 2015). The current study looks at the relationship of alcohol inebriation with suicidal ideation upon sobriety. It is possible that patients may be at increased risk for suicide *while they are intoxicated*. The current study did not address the ability to predict suicide upon re-inebriation. However, the proximate concern of the emergency provider and behavioral health specialists is to establish safety upon discharge.

The study results should be interpreted in the context of its strength and limitations. This was a retrospective study, necessitated by the fact that ethical consent is very challenging for patients who are intoxicated and/or have a potentially serious psychiatric condition, both of which constitute important vulnerable populations. The study avoided confounding by the COVID-19 pandemic by using data prior to its onset.

In the current study, no specific scale or instrument was used to determine a final diagnosis of suicidality. Various instruments have been designed to evaluate the risk of suicide and safe disposition, and the American College of Emergency Physicians (ACEP) clinical policy criticize these scales as being ineffective at predicting the risk of self-harm or death (Mullinax et al 2018).

Dual diagnosis is a term used to describe those who have both a mental health diagnosis and a co-occurring substance use disorder. This is a frequently seen subset of patients in emergency health settings. Thorough assessment and treatment of each diagnosis is accepted best practice for these patients (Rodriguez-Cintas et al 2017) (Szerman N et al 2012). For this study, we acknowledge that many of the participants are represented in this category. We have chosen to focus solely on examining any relationship between intoxication and suicidality irrespective of “dual diagnosis.”

No inter-rater reliability for the final diagnosis of suicidality was possible due to the fact that the study was entirely retrospective. The authors acknowledge that there may be some variability between the diagnosticians who determined the final diagnosis of SI. No attempt was made to evaluate the data using a block analysis based on individual practitioners.

The Department of Transportation’s Appropriations Act for FY2001 made federal highway construction funds dependent on states enacting laws prohibiting driving with 0.08 g/dL (80 mg/dL) or greater blood alcohol concentration (BAC) (US Department of Transportation, 2001). Although sobriety has been defined in terms of these specific blood levels, they may not correlate with clinical sobriety in all individuals. It is known that alcohol tolerance can impact cognitive ability and mechanical performance (NIAAA 1995) (Tabakoff et al 1986). For example, chronic alcohol use is known to result in increased tolerance in some individuals, and these individuals may be clinically sober at higher levels of serum alcohol (Roberts et al 2010).

The presence of co-ingestants was documented as part of the data abstraction process. However, no analysis of association with the major outcome measure was possible for specific agents or a combination of agents due to the size of the study population. Also, it is impossible to determine if patients with positive urine drug screens are under the influence of that particular agent at the time of the test. Many agents are known to be detectable by qualitative methods long after their effects are clinically manifested. Future research should evaluate the impact of various individual agents on SI, perhaps using a similar methodology as the current study. This may require the use of quantitative methods to correlate more closely with intoxication.

## Conclusion

In this study of pre-COVID-19 patients presenting to an emergency department with suicidal ideation, intoxication with alcohol was clearly associated with a lower level of reliability as measured by behavioral professional specialist evaluation upon sobriety. Prior to sobriety, the accuracy of a determination of active suicidality can be expected to be low. This suggests that it is prudent to await sobriety prior to the definitive assessment of patients with a risk of self-harm.

## Data Availability

All data produced in the present study are available upon reasonable request to the authors

